# A distinct T cell receptor signature associates with cardiac outcome in myocardial infarction patients

**DOI:** 10.1101/2023.11.23.23298725

**Authors:** Kenz Le Gouge, DiyaaElDin Ashour, Margarete Heinrichs, Paul Stys, Pierre Barennes, Verena Stangl, Lavinia Rech, Gerald Hoefler, Karl Kashofer, Tobias Gassenmaier, Valerie Boivin-Jahns, Roland Jahns, Ulrich Hofmann, Dominik Schmitt, Anna Frey, Stefan Störk, Stefan Frantz, Peter P. Rainer, Gustavo Campos Ramos, Encarnita Mariotti-Ferrandiz

**Affiliations:** Sorbonne Université, INSERM, UMRS959, Immunology-Immunopathology-Immunotherapy (i3), Paris, France; Department of Internal Medicine I, University Hospital Würzburg, Würzburg, Germany; Comprehensive Heart Failure Center, University Hospital Würzburg, Würzburg, Germany; Diagnostic and Research Institute of Pathology, Medical University of Graz; Division of Cardiology, Medical University of Graz, Graz, Austria; Department of Diagnostic and interventional Radiology, University Hospital of Würzburg, Germany; Institute of Pharmacology and Toxicology, University of Würzburg, Würzburg, Germany; Interdisciplinary Bank of Biomaterials and Data Würzburg (IBDW), University Hospital of Würzburg, Würzburg, Germany; BioTechMed Graz, Graz, Austria; St. Johann in Tirol General Hospital, St. Johann in Tirol, Austria; Institut Universitaire de France (IUF)

**Keywords:** Myocardial infarction, ejection fraction, T cell, TCR, lymphocytes

## Abstract

Myocardial infarction (MI) is associated with an inflammatory process mainly attributed to innate immune components. Very recently, the role of T-cells in both inflammation and healing has been suggested through various human and mouse studies. Previous studies showed that CD4+ and CD8+ T cells affect post-MI repair but did not investigate how to leverage T cell biology to predict post-MI outcomes in patients. For instance, the antigenic trigger of T-cells is still unknown in human. Indeed, others and we identified T-cell specific for myosin infiltrating the myocardium in mouse models of MI, recent studies identified expanded clones in human myocardium, altogether suggesting a tissue-specific T-cell activation. However, it is still unclear how acute post-MI immune responses shape long-term cardiac functional outcomes in individual patients. In this study, we analyzed the role of T-cell in predicting post-MI repair by analyzing the T-cell receptor (TCR) repertoire. Indeed, the TCR repertoire is now considered as a marker of the clinical status of individuals. Previous studies in infectious but also autoimmune contexts showed the potential of the TCR repertoire to predict the disease. Therefore, assessing the dynamic changes in global TCR repertoires may provide valuable information about the antigen-specific immune responses underlying post-MI healing. In our study, we carefully selected patients that suffered from MI on a prospective cohort. The TCR repertoire has been analyzed by next generation sequencing at the index hospitalization with the aim to identify features predict of their healing outcome assessed at 12 months post-MI. While no major variations have been found in diversity of TCR gene usage, we identified unique TCR signatures predicting one-year cardiac functional outcomes. Our result enables early immune-based risk stratification of MI patients and calls for larger studies to develop novel predictive biomarkers and possibly new therapeutics.

## Main text

Inflammatory processes govern post-myocardial infarction (MI) healing and remodelling. Yet, we cannot predict how acute post-MI immune responses shape long-term cardiac functional outcomes in individual patients. Previous studies showed that CD4^+^ and CD8^+^ T cells are key actors of the post-MI repair regulation(1, 2) but did not investigate how they could be used as predictors of the cardiac outcome. Here, we sought to identify T cell receptor (TCR) signatures predicting cardiac functional outcomes in a well-characterized MI patient cohort(3).

TCRs are heterodimeric antigen-specific receptors expressed by T cells, generated through somatic recombination of multiple gene segments in a process that generates a potential repertoire of 10^19^ unique receptors(4). Thus, we hypothesised that analysing TCR repertoires in patients after MI could provide valuable information on the individual immunological status underlying post-MI healing outcomes.

First, we selected acute ST-elevation MI-patients from the ETiCS cohort (**<u>E</u>**tiology, **<u>Ti</u>**tre-Course, and effect of autoimmunity on **<u>S</u>**urvival study(3), Würzburg arm) exhibiting reduced left ventricular ejection fraction (LVEF) assessed by cardiac magnetic resonance (cMRI) on day 4 post-MI (LVEF <50%, 54/150 patients). Amongst those, we further selected patients with complete serial cMRI scans at baseline and 12 months of follow-up (FU) (38/54 patients) (**Figure 1A**). Next, we stratified these patients into “good” versus “poor” healers based on a priori defined cMRI criteria(5)). In brief, patients showing a ΔLVEF <13% between baseline and follow-up (FUP) were defined as “poor healers” (25/38), whereas those showing greater improvement (ΔLVEF >13%) were considered “good healers” (13/38) (**Figure 1B**). In addition to lower ΔLVEF values (P<0.0001), poor healers had significantly greater Δ end-systolic volumes (ESV) (P<0.0001), whereas Δ end-diastolic volumes (EDV) did not differ. Age, BMI, infarct size, and routine blood biomarkers were similar in poor and good healers at baseline (**Figure 1B**). After defining groups of MI-patients with diverging healing phenotypes, we extracted RNA of cryopreserved whole blood samples that were collected at hospital admission from all 25 poor and 13 good healers (Nucleospin RNA blood kit, Macherey-Nagel, Düren, Germany). Due to technical limitations inherent to the processing of archived biomaterial, we obtained RNA with sufficient quality from 19 poor and 9 good healers, which were then used for RT-PCR, amplification of TCR alpha and beta chains (TRA and TRB respectively), library preparation and sequencing(6).

**Figure 1:**
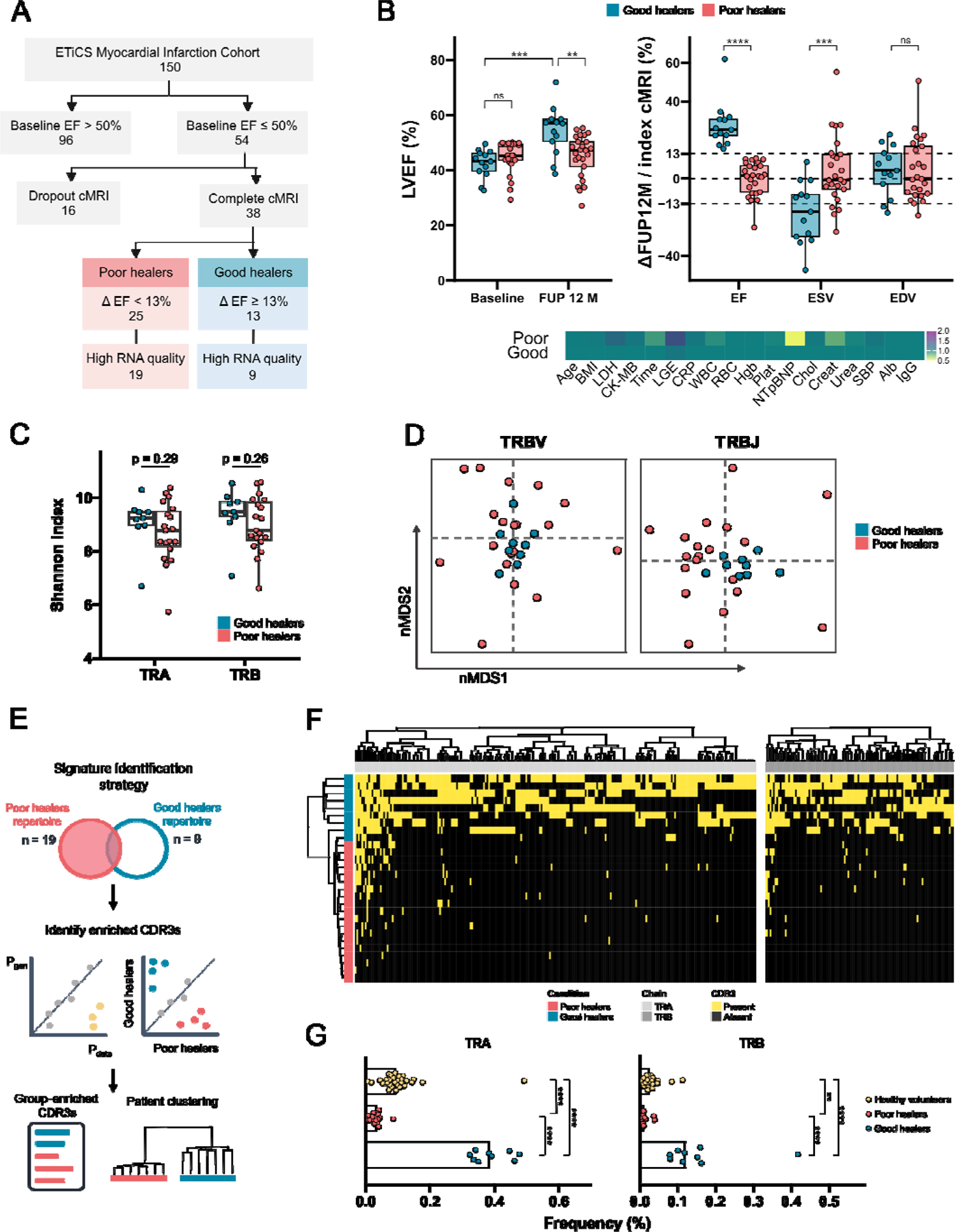
A distinct T cell receptor signature associates with cardiac functional outcomes in myocardial infarction patients. **(A)** Patient selection and stratification into good vs poor healers based on cMRI findings. The number of patients is indicated within the boxes (**B**) cMRI findings, including LVEF at baseline and FUP 12 months in good vs poor healers, ΔLVEF (%), ΔESV and ΔEDV (mL), and distribution of confounding factors between groups (BMI: body-mass index; LDH: lactate dehydrogenase; Time: time from pain to intervention; LGE: infarct size assessed by late gadolinium enhancement, CRP: C-reactive protein, WBC: white blood cell count; RBC: red blood cell count, Hgb: hemoglobin; Plat: platelets count; NTproBNP; N-terminal pro-brain natriuretic peptide; Chol: cholesterol; Creat: creatinine; SBP: systolic blood pressure; Alb: albumin. Values are expressed as fold change from the average levels found in the “good healer” group). (**C**) Shannon index based on clone counts from aligned TCR samples for the alpha and beta chain. P-value was computed using Wilcoxon U-test. (**D**) Non-metric dimensional scaling of the TRB variable (TRBV) and joining (TRBJ) gene usage frequencies. (**E**) Strategy used to identify Complementarity-determining region 3 (CDR3, antigen-binding domain) signature. (**F**) Heatmap of the 348 CDR3 signature found in patients. CDR3, by column, were clustered independently by chain. (**G**) Sample’s cumulative frequencies (in %) of the CDR3 from the signature. Healthy volunteers are depicted in yellow.

To evaluate potential global changes in the TCR repertoires found in blood sampled at baseline, we first measured the diversity of TCRs in each group by computing the Shannon entropy and the gene usage frequency (**Figure 1C-D**), two canonical measures of immune repertoire diversity(7). These basic analyses did not reveal any significant differences between groups. We then hypothesised that distinct TCR repertoire composition may explain healing outcomes. Thus, we devised a refined strategy to identify a set of TCRs that effectively differentiates between cardiac functional outcomes (**Figure 1E**). Accordingly, using statistical modelling from Pogorely *et al*.(8) (**Figure 1E**, panel 1), we computed the probability of each TCR being present in the dataset (Pdata) and identified TCRs that were statistically enriched compared to their expected generation (Pgen). Additionally, we searched for TCRs that were differentially represented between good and bad healers(9) (**Figure 1E**, panel 2). Finally, we selected TCRs that met both criteria: being more represented in one group than the other and having a statistically higher probability of being present in the dataset than expected (**Figure 1E**, panel 3). As a result, we discovered a signature of 348 unique TCRs (**Figure 1F**). Hierarchical clustering based on the presence/absence of this signature allowed a separation of good from poor healers (cluster purity = 0.93). Interestingly, this signature comprising 237 unique TRA and 111 unique TRB was primarily present among the good rather than the poor healers. We then assessed the relevance of the signature by comparing its cumulative frequency in the ETiCS patients versus a group of healthy subjects (n = 27). **Figure H** shows that the cumulative frequency of the signature is significantly higher in good healers when compared with poor healers and healthy volunteers (p<0.001 for both TRA and TRB chains). Moreover, the TCR signature was more enriched in healthy subjects when compared with poor healers (p<0.001 for TRA and p<0.01 for TRB), suggesting that the clones found in the signature are specifically depleted in MI patients with poor cardiac functional recovery (**Figure 1G**).

In summary, we identified a distinct TCR signature associated with cMRI-assessed post-MI cardiac healing outcomes. Specifically, we found that MI-patients showing a substantial post-MI LVEF improvement were enriched for a set of 348 unique TCRs in the peripheral blood already at baseline (hospitalization). These exploratory findings provide clinical evidence that the individual TCR profile at early stages post-MI contributes to long-term healing outcomes in humans. This supports the notion that modulation of T cell responses may eventually help to improve post-MI remodelling(10) and further confirms that the circulating T cell compartment is impacted by the ongoing cardiac repair. Moreover, the present findings provide a compelling example on the value of immune-based diagnostics in cardiology. Still, our study has some important limitations. First, it was a rather small - though well stratified and characterized - retrospective patient collective. Moreover, since we analysed cryopreserved whole blood samples, we were unable to phenotype T cell subsets that might have accounted for the identified TCR signature. Furthermore, as it is currently not possible to infer which antigens are recognised by the TCRs comprised in this distinct signature, the mechanisms underlying their association with the cardiac functional outcomes remain unknown. Still, our findings underline the potential predictive value of TCR repertoire analyses in MI and might encourage further mechanistic investigations and confirmation in larger patient cohorts.

## Author contribution

RJ, VJ, DS, AA, SS recruited the patients for this study and coordinated the clinical study; TG analysed the cMRI images, MH, DA, LV, VS, PPR and GCR retrospectively stratified and selected samples; DA, KLG, PS, PB, and EMF processed the RNA samples, developed bioinformatic tools and analysed the TCR-seq data. DA, MH, KLG, GH, UH, SF, PPR, EMF, GCR made substantial contributions to the conception and design of the present work. KLG, DA, PPR, EMF, and GCR drafted the manuscript. PPR, EMF, and GCR supervised this study. All co-authors critically revised it, made significant intellectual contribution, and approved the manuscript in its final version to be published.

## Data Availability

All data produced in the present study are available upon reasonable request to the authors.

## Acknowledgements

This work benefited from equipment and services from the iGenSeq core facility at Institut du Cerveau (ICM, Paris, France) for all the TCR data production.

## List of non-standard abbreviations

MRI: cardiac magnetic resonance imaging

ESV: end-systolic volume

EDV: end-diastolic volume

FUP: follow-up visit

HF: Heart failure

LVEF: left-ventricular ejection fraction

MI: myocardial infarction

STEMI: ST-elevation myocardial infarction

TCR: T cell receptor

TRA: T cell receptor, alpha chain

TRB: T cell receptor, beta chain

